# Genetic evidence of cross-border *Plasmodium vivax* spread in a malaria pre-elimination region of South Asia

**DOI:** 10.64898/2026.02.02.26345415

**Authors:** Anjana Rai, Edwin Sutanto, Prakash Ghimire, Sonam Wangchuk, Tobgyel Drukpa, Mohammad Shafiul Alam, Najia Ghanchi, Awab Gulam Rahim, Angela Rumaseb, Hidayat Trimarsanto, Kian Soon Hoon, Nabaraj Adhikari, Sanjib Adhikari, Megha Raj Banjara, Komal Raj Rijal, Roshan Nepal, Ramesh Regmi, Bushra Qurashi, Shan-e-Zehra Zaidi, Mohammad Asim Beg, Benedikt Ley, Ric N Price, Kamala Thriemer, Sarah Auburn

## Abstract

**Background:** *Plasmodium vivax* is the predominant cause of malaria in South Asia. *P. vivax* cases have fallen over the past decade, but cross-border transmission remains a major challenge to elimination. Genetic data can generate valuable insights into transmission; however, until now, only low-resolution data have been available from Nepal, Bhutan and Bangladesh. We piloted high-resolution genotyping using a new microhaplotype (multiallelic) assay to monitor *P. vivax* transmission across borders.

**Methods:** Genotyping was conducted using the 93-microhaplotype vivaxGEN panel on *P. vivax* parasites collected from patients enrolled in clinical trials in Bangladesh, Bhutan, and Nepal between 2013 and 2023. These data were compared with open-access microhaplotype and genomic data derived from Afghanistan, India, and Pakistan between 2014 and 2024. Polyclonality and relatedness (identity by descent (IBD)) were determined within and between countries.

**Results:** High-quality genotyping data were generated for Nepal (n = 19), Bhutan (n = 27), and Bangladesh (n = 35); comparative data were sourced from Afghanistan (n = 159), India (n = 24), and Pakistan (n = 213). Overall, 29.6% (47/159) of isolates from Afghanistan and 20.2% (43/213) from Pakistan had polyclonal infections, whereas all parasites from Bhutan, Nepal and Bangladesh were monoclonal, suggesting lower superinfection. Country-wide IBD analyses revealed three genetic clusters partitioning Bangladesh and Bhutan (partial) from the remaining countries. There were two sub-populations in Bhutan, which separated local and cross-border imported cases.

**Conclusions:** Our results highlight the use of regional high-resolution genetic data to enhance monitoring of transmission intensity and cross-border importations.

## Introduction

Malaria remains a major public health threat in South Asia, with more than 6 million cases reported in 2024; *Plasmodium vivax* accounted for approximately 60% of these cases, contributing substantially to the socio-economic burden. [1]. India, Pakistan and Afghanistan remain highly endemic, accounting for almost 70% of all *P. vivax* malaria globally, whereas over the last decade, *P. vivax* cases have declined significantly in Nepal, Bhutan and Bangladesh [1]. Cross-border movement is frequent in South Asia, sustaining the reintroduction of *P. vivax* parasites into elimination areas such as the borders of Bhutan and Nepal. In Bhutan, there have been no local malaria cases since 2022; however, the southern border with India remains highly porous, with recent cases being imported, emphasising the ongoing risk of parasite importation and resurgence [1, 2]. Nepal also shares a border with India to the east, south, and west, exposing the country to importation of parasites from malaria-endemic regions. Extensive cross-border movement of migrant workers returning from India to Nepal resulted in a high number of imported malaria cases; in 2023, these accounted for 98% of all *P. vivax* cases [1, 3]. Monitoring *P. vivax* transmission within and across borders is challenging owing in part to the biology of this species.

*P. vivax* can form dormant liver stages (hypnozoites), which reactivate weeks to months after initial infection causing recurrent blood-stage infections that facilitate ongoing transmission and a propensity for resurgence. Individuals with asymptomatic *P. vivax* infections act as the main reservoirs for transmission [4]. Furthermore, recent studies reveal that a substantial parasite biomass accumulates in the spleen, forming another hidden reservoir that likely sustains local transmission and cross-border spread [5]. Amidst this complexity, genetic data has potential to inform on parasite transmission dynamics.

Prior to our study, the genetic diversity and associated insights on *P. vivax* transmission and cross-border spread in South Asia remained poorly characterised. Previous investigations in Nepal, Bhutan and Bangladesh used genotyping at panels of up to 10 microsatellites to assess local *P. vivax* diversity [6-8]. While microsatellites are highly diverse, their genotyping costs are high, which limits panel size and associated resolution for detecting fine-scale population structure and relatedness [9, 10]. It is also challenging to collate microsatellite data from different studies unless consensus control samples and variant calling methods are applied prospectively [11-13]. This limits cross-country comparisons.

In this study, we present the first high-resolution *P. vivax* genetic datasets from Bhutan, Nepal and Bangladesh using a recently established 93-microhaplotype panel [13]. When these datasets were combined with existing data from three highly endemic countries in the region, the high-resolution marker panel permitted measures of identity-by-descent (IBD) to explore parasite-relatedness within and between countries. Our findings highlight the potential of this approach to track cross-border spread of *P. vivax* and provide important baseline data for ongoing surveillance of *P. vivax* transmission and spread across South Asia.

## Material and Methods

### Patient samples from new datasets

Parasite isolates were collected from patients enrolled in clinical studies conducted between 2013 and 2024 in five South Asian countries (Supplementary Table 1; Supplementary Figure 1). Samples from Nepal were collected from a clinical trial in Sudurpashchim Province, between 2021 and 2023, evaluating the safety and efficacy of a low-dose short-course primaquine treatment (3.5mg/kg total dose given over 7 days) in glucose-6-phosphate dehydrogenase (G6PD) normal patients infected with *P. vivax* and *P. falciparum* (ClinicalTrials.gov ID: NCT04079621). Samples from Bhutan were collected during a therapeutic efficacy survey conducted from 2013 to 2015 in Sarpang, Gelephu, Trongsa, Wangdiphodrang, Trongsa, Pemagatshel and Samdrupjongkhar districts [6]. Patients enrolled in the study were categorised into two groups based on nationality: Bhutanese nationals and non-nationals, to examine whether 93-microhaplotype assay can capture genetic separation between locally acquired and cross-border imported *P. vivax* infections. Samples from Bangladesh were collected during a prospective observational therapeutic efficacy study in the Chittagong Hill Tracts conducted between 2014 and 2015 [14].

DNA was extracted from blood samples using the Qiagen QIAamp DNA Blood Mini or Midi Kit, according to the manufacturer’s instructions (Qiagen, Hilden, Germany). *Plasmodium* species were confirmed by PCR as previously described by Padley *et al*. [15] with a modification suggested by Boonma *et al*. [16] that each species is tested in a singleplex assay.

### Existing data for comparative analysis

Amplicon sequencing and whole-genome sequencing (WGS) data from high-endemic countries in the region were included for comparative analysis. Amplicon sequencing data for Afghanistan were sourced from a previous study, which included samples collected during a clinical trial conducted from 2014 to 2017 in Jalalabad and Laghman (ClinicalTrials.gov ID: NCT01814683) [13, 17]. Amplicon sequencing data from Pakistan were generated using samples from a clinical trial (ClinicalTrials.gov ID: NCT04411836) conducted between 2023 and 2024 in Karachi and Thatta [18]. The samples from both clinical trials were collected from patients presenting with clinical symptoms, with *P. vivax* parasitaemia confirmed by blood film microscopy. The amplicon sequencing data from Afghanistan and Pakistan were generated using the same library preparation methods applied to the new datasets from Bangladesh, Bhutan and Nepal for the vivaxGEN 93-microhaplotype panel [13]. Genetic data from India were obtained from an open-access whole-genome sequencing (WGS) dataset generated using standard Illumina library preparation on genomic DNA samples, with Illumina sequencing generating 150bp paired-end reads [19].

### Selective whole genome amplification

*P. vivax* selective whole-genome amplification (sWGA) was performed on samples from populations where pilot genotyping demonstrated low genotyping success in genomic DNA (gDNA) samples. sWGA was performed using two previously published primer sets, pvset1920 and pvset1 [20]. Amplifications were conducted separately with each primer set, and a pool of each sWGA products equal volume was also prepared for downstream library preparation.

### Amplicon sequencing

Genomic DNA (gDNA) or sWGA product were used as input for library preparation using the vivaxGEN microhaplotype panel as previously described [13]. Briefly, libraries were prepared using a rhAmpSeq kit (Integrated DNA Technologies) with 5 μL template in 10 μL reactions to generate amplicons at the custom primer pool, followed by indexing. The resulting libraries were paired-end sequenced in multiplexes of up to 192 samples on an Illumina MiniSeq platform using the MiniSeq Mid Output kit (300 cycles).

### Variant calling

Variant calling on the amplicon data was performed using the DADA2-based vivaxGEN MicroHaps pipeline (commit 359d042, https://github.com/vivaxgen/MicroHaps). The data were further processed to remove insertions and deletions (indels), and alleles with less than 2% proportion-wise or 5 read-pairs per sample. In the WGS data, SNP variants were initially called, requiring at least 5 reads to call an allele, with heterozygous calls if the minor allele had at least 2 reads and contributed to >=5% of the total reads. Indel variants were removed, and then only microhaplotypes with no missing or heterozygous SNPs were retained. The amplicon and WGS data were combined and then samples with at least 60% (55/93) microhaplotype markers genotyped were included in downstream analyses.

### Population genetics analysis

Within-host diversity was measured using the effective Multiplicity of Infection (eMOI) metric, determined by MOIRE software (v3.5.0) [21]. The following parameters were used for MOIRE: burnin=1000; pt_chains=seq(0.5, 1, length.out=40); samples_per_chain=1000; r_alpha=3; r_beta=3. eMOI is a metric of diversity that incorporates MOI and relatedness. Relatedness between samples was measured using Dcifer (v1.2.1) [22]. Pairwise identity by descent (IBD) was estimated in Dcifer with the assumption that paired strains between samples have the same relatedness. The total IBD between samples were then scaled by the minimum MOI between the two samples to obtain the overall relatedness. Network plots showing relatedness between different samples were created using ggnetwork (v0.5.13) [23].

The following analyses were conducted on a subset of isolates that were monoclonal. Isolates are categorised as monoclonal if they had <50% polyclonal probability, generated by MOIRE estimates. Microhaplotype marker diversity and effective cardinality in the different countries was undertaken using paneljudge [24], with marker 64721 removed from paneljudge analysis. To confirm the population structure observed in Dcifer analysis, we used structure (v2.3.4) [25] with the following parameters: 10,000 burnin; 20,000 numreps; and 1 ploidy. Other parameters were left as default. We tested different K values (1 to 9) and performed 20 repeats for each K with randomised seeds. Sorting and plotting of the structure results were done using CLUMPAK [26]. The optimal number of clusters was calculated using the delta K method [27].

### Ethics approval

Written, informed consent was provided by all patients or a parent or guardian. Ethical approvals for all studies included the use of blood samples for parasite genotyping and were provided by the Menzies School of Health Research Human Research Ethics Committee (HREC) of the Northern Territory Department of Health and local ethics committees as listed in Supplementary Table 3.

## Results

### High-quality microhaplotype data generated

Genotyping data were successfully generated for 96.4% (27/28) of samples from Bhutan, 89.7% (35/39) from Bangladesh, and 86.4% (19/22) from Nepal. Open-access data were obtained from 159 samples from Afghanistan, 24 from India and 213 from Pakistan. In total, high-quality amplicon sequencing data were generated on 453 independent samples, with additional WGS-derived data on 24 samples. High marker diversity was confirmed in all populations, providing high genetic resolution for diversity and structure analysis (Supplementary Figure 2). A summary of the available patient demographics for the successfully genotyped samples is presented in Supplementary Table 2, and line-level sample details are presented in Supplementary Data 1. Variation was observed amongst the populations in gender, but in all populations, majority were male (range 60-89%) (Supplementary Table 2). Aside from Afghanistan and Bangladesh, where a high proportion of individuals were in the 5–15-year-old category (34-73%), >80% of patients were in the >15-year-old category.

### Infrequent superinfection in the low-endemic populations

Polyclonality and within-host infection diversity can be used as a proxy to transmission intensity, with greater diversity generally observed in regions with higher transmission and associated superinfection and co-transmission [11]. For polyclonal estimation and within-host diversity analysis, the 24 WGS-derived cases from India were excluded from analysis as the cases were pre-selected to be monoclonal. Amongst 453 independent samples with amplicon-based sequencing data, polyclonal infections were observed in 29.6% (47/159) of isolates from Afghanistan and 20.2% (43/213) from Pakistan (Figure 1a). All 35 isolates from Bangladesh, 27 from Bhutan and 19 from Nepal were monoclonal. Within-host infection complexity was assessed using the effective multiplicity of infection (eMOI). In line with the polyclonality data, the lowest eMOI distributions were observed in Bangladesh, Bhutan and Nepal (mean eMOI = 1), suggesting low superinfection consistent with low endemic settings (Figure 1b). In contrast, higher eMOI distributions were observed in Afghanistan (mean eMOI = 1.22) and Pakistan (mean eMOI = 1.11), suggesting a higher incidence of superinfection and co-transmission (Figure 1b).

**Figure 1.**
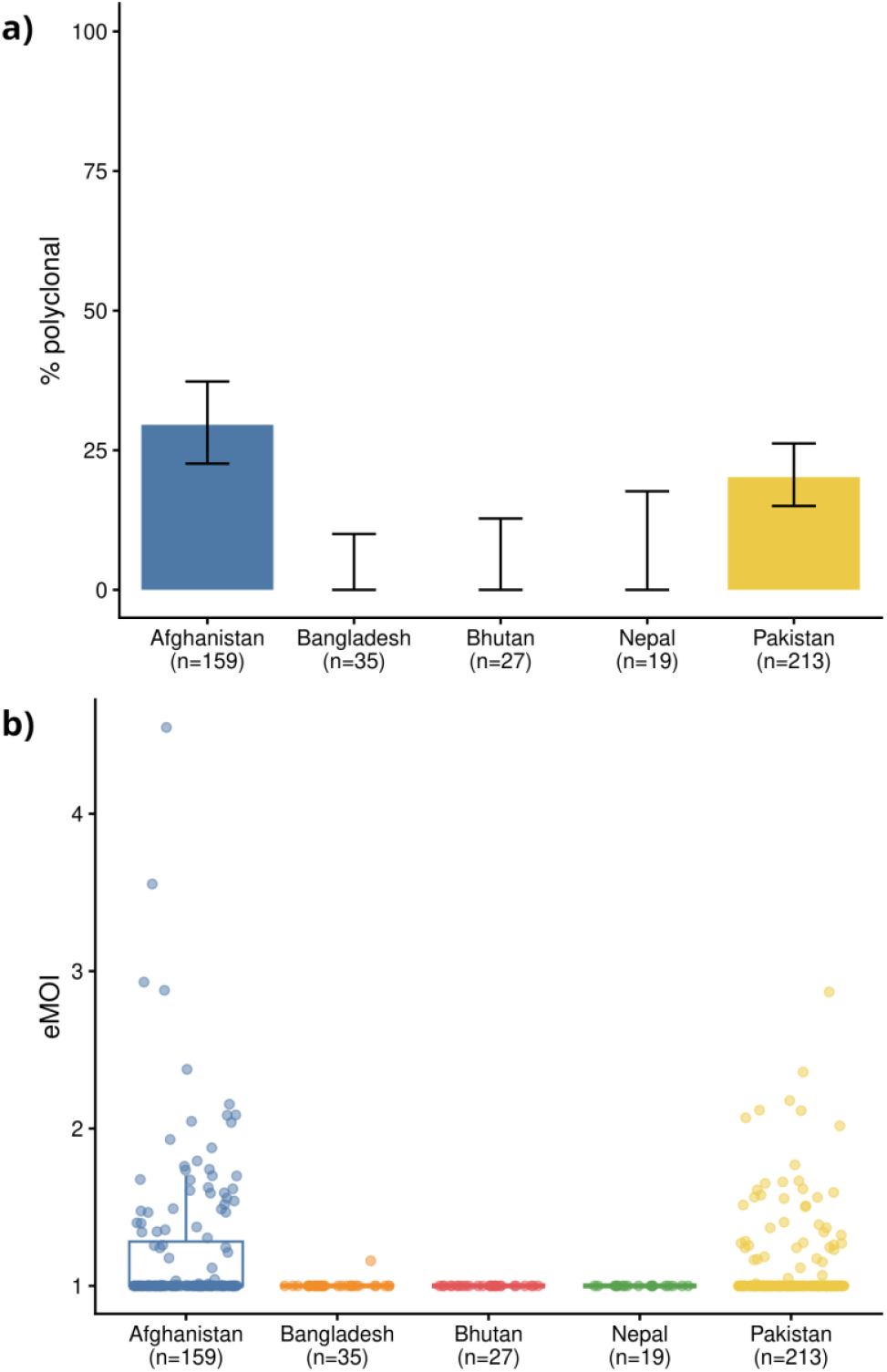
Low within-host infection diversity in low-endemic countries. Panel a) presents the percentage of polyclonal infections, and panel b) presents the estimated multiplicity of infection (eMOI) distribution by country. Afghanistan and Pakistan have higher annual parasite incidence than the other populations and accordingly higher polyclonality and eMOI. India was excluded owing to the biased pre-selection for low-complexity genomic samples to enable accurate microhaplotype calling.

### Evidence of a large cross-border reservoir

Analysis of IBD was conducted at the regional level to inform on the relatedness between parasites from different countries. There were three distinct infection clusters in the IBD-network plot with IBD threshold of 6% (Figure 2a). The samples from Afghanistan, India, Pakistan and Nepal formed one large cluster, whilst the samples from Bangladesh formed a distinct cluster. The samples from Bhutan were divided into two sub-populations: one group formed its own cluster, while the other group clustered together with the multi-country cluster. Closer inspection of the Bhutanese sub-populations by patient nationality revealed that the distinct group largely comprised Bhutanese nationals, whilst the multi-country group largely comprised non-nationals, including Indian nationals presenting with clinical illness in Bhutan (Figure 2a).

**Figure 2.**
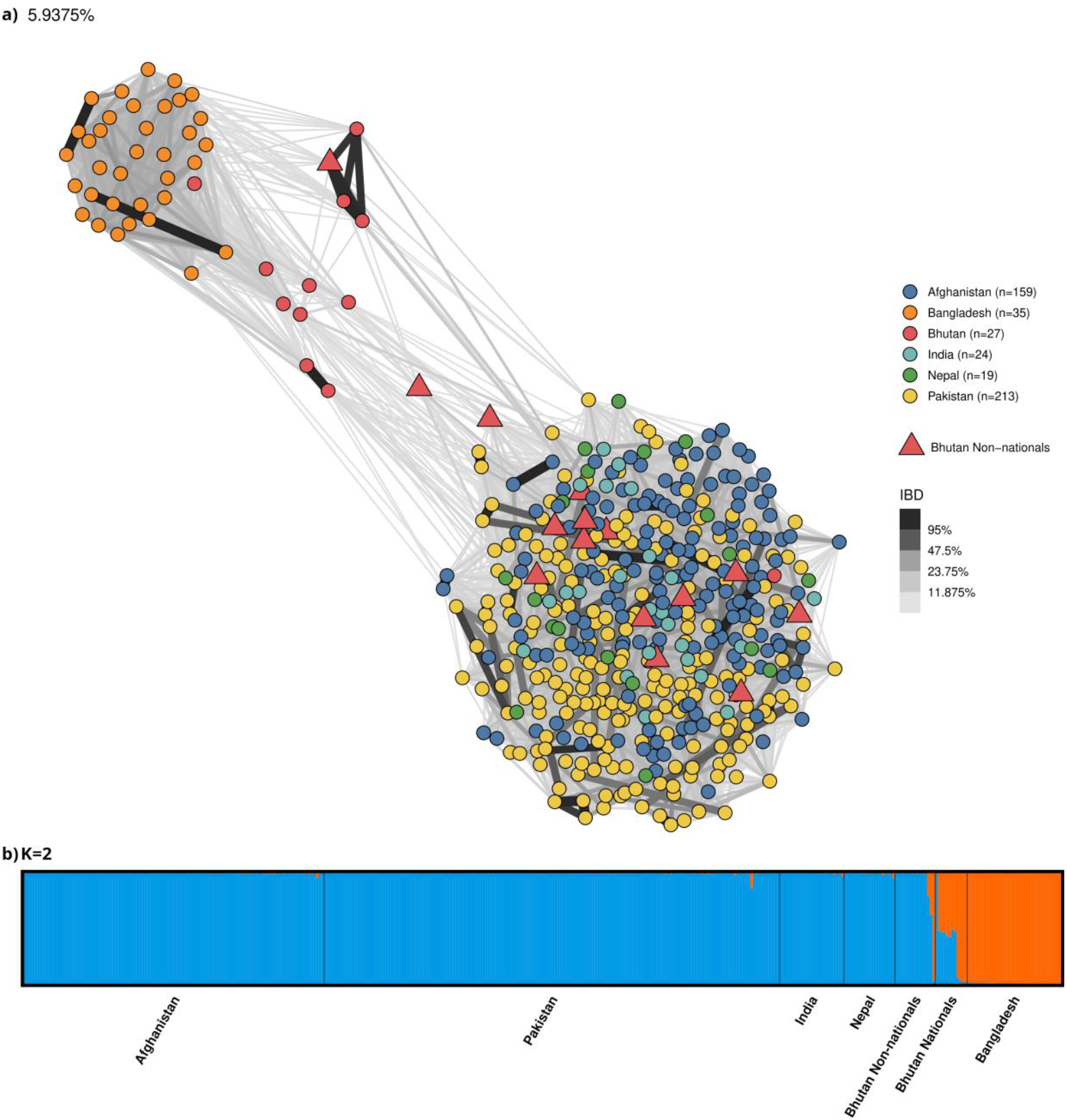
Regional population structure reveals cross-border transmission. Panel a) presents an identity-by-descent-based (IBD-based) network illustrating connectivity between infections from Bhutan, Bhutan non-nationals (are of Indian origin, represented by a triangle shape), Bangladesh, Nepal, Afghanistan, India, and Pakistan. Each shape represents an infection and is colour-coded by sites. For each country, connectivity, illustrated by a grey to thick black connecting line, is presented at the IBD threshold of ≥6% (thin grey lines) to ≥95% (thick black lines). Panel b) presents STRUCTURE-based population grouping. Across the six countries. The optimal number of clusters was determined using the delta K method, which showed the highest value at K = 2. Each bar represents an individual isolate with colour-coding reflecting the ancestry to each of the two K groups (K1 in blue and K2 in orange). The Bangladesh isolates formed a distinct cluster (K2), while Afghanistan, Pakistan, India, Nepal, and Bhutan non-national isolates clustered together (K1). In contrast, the Bhutanese national isolates showed mixed ancestry to K1 and K2 or predominant K2 ancestry. Analysis was conducted on 477 and 387 independent infections for Dcifer and STRUCTURE, respectively.

Further investigation of population structure in the regional dataset was conducted using STRUCTURE software. The optimal number of clusters (K populations) [27] was identified as 2 (Supplementary Figure 3). 89.9% (337/375) of isolates from Afghanistan, Bhutanese non-nationals, India, Nepal and Pakistan had major ancestry to a large multi-country group (K1), whereas the 58.3% (7/12) of Bhutanese nationals showed mixed ancestry to both groups (Figure 2b). At K = 2, all (35) Bangladesh isolates had a majority (>80% ancestry) to a distinct cluster (K2) from isolates from the other South Asian populations (Figure 2b).

### Variation in inbreeding between sites

Analysis of within-country IBD was conducted to inform on the relatedness between parasites within countries. The samples from Bangladesh showed high relatedness (median IBD = 19.2%) compared to those from other countries included in the analysis. In contrast, Bhutan Nationals (median IBD = 1.5%), Bhutan Non-nationals (median IBD = 0%), and Nepal (median IBD = 2%) showed similar relatedness to those in the higher endemic settings (Figure 3).

**Figure 3.**
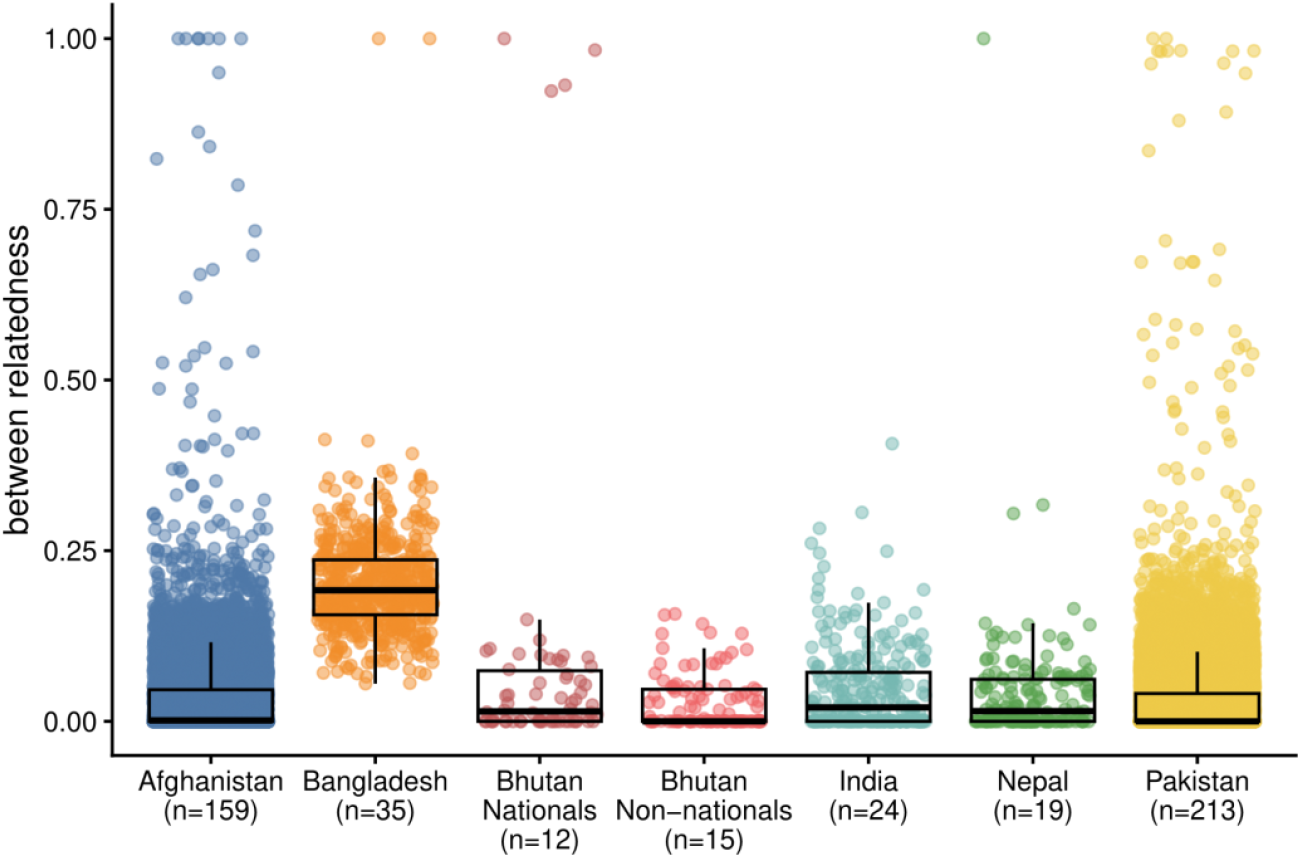
Variation in between-host infection relatedness by country. The boxplots present the distribution of the identity-by-descent (IBD) values for each country, providing the median and interquartile ranges.

### Demonstrated strength of high-versus low-resolution markers to capture IBD and separate local vs imported cases in Bhutan

Previous analyses using a panel of 9 microsatellites did not reveal genetic separation between locally acquired and cross-border imported *P. vivax* infections in Bhutan [6]. We explored whether the separation observed with the 93-microhaplotype assay was due to the panels capacity to capture IBD accurately. Paneljudge software was used to assess the diversity of the two marker panels and their accuracy in predicting different levels of IBD between sample pairs in Bhutan. Although the microsatellites had a higher average marker diversity and effective cardinality compared to the microhaplotypes (Supplementary Figure 4a and 4b), the relative mean square error (RMSE) of the microsatellites was up to two-fold higher than for the microhaplotypes (Figure 4a). IBD networks illustrated the relative limitation of the microsatellite panel, with almost all samples (national and non-national) showing relatedness at ∼12% (Figure 4b). In contrast, the microhaplotypes showed greater discrimination of the Bhutanese non-nationals (related at ∼3%), indicating the panels greater ability to resolve low relatedness (Figure 4b). Overall, the data suggested that 93 microhaplotypes could estimate IBD with significantly greater accuracy than the 9-microsatellite assay. Even without reference data from other countries, the 93 microhaplotype panel achieved greater differentiation of local versus imported infections in Bhutan.

**Figure 4.**
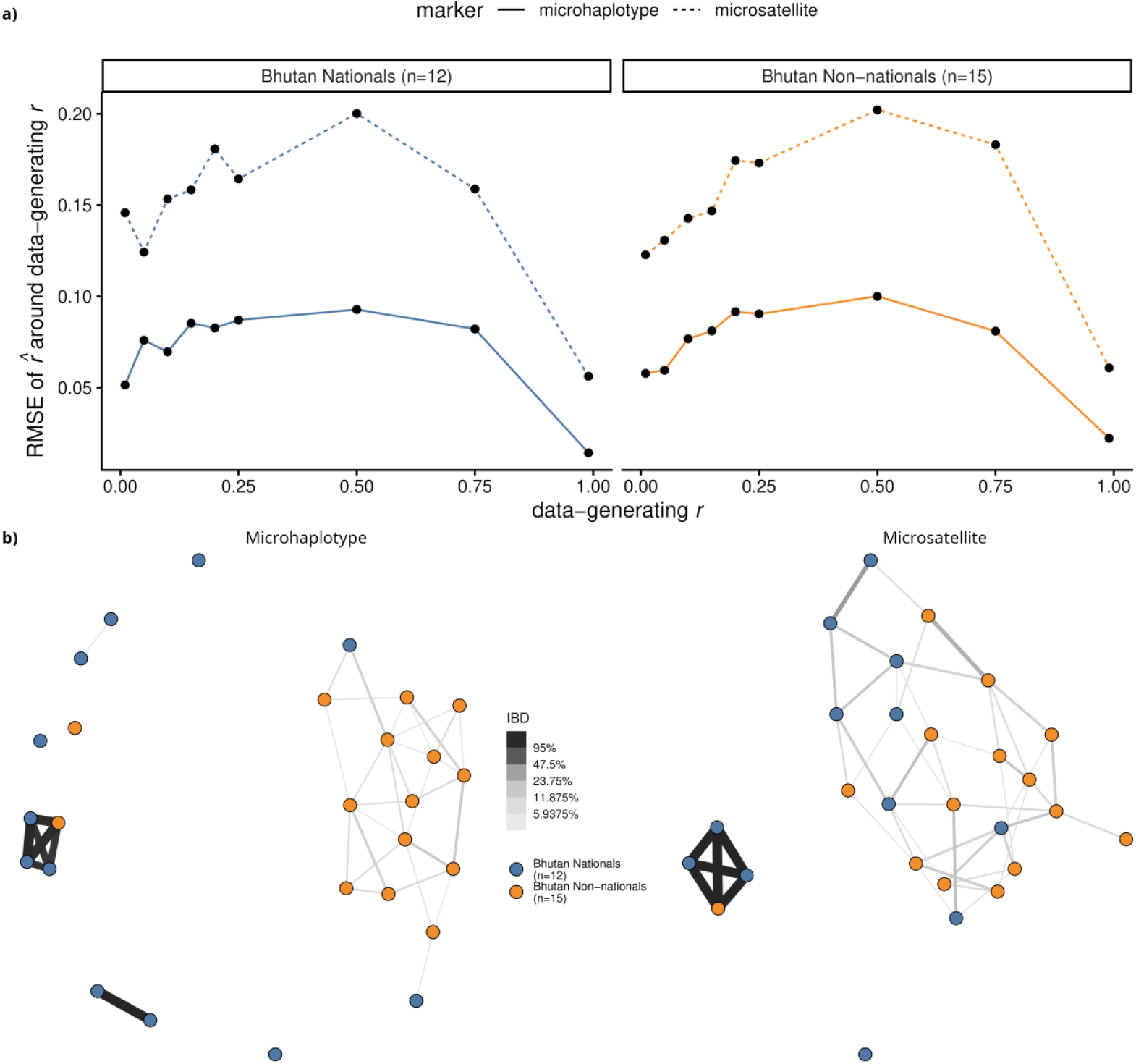
Higher IBD estimation accuracy in Bhutan using 93 microhaplotypes versus 9 microsatellites. Panel a) presents the relative mean square error (RMSE) in estimation of the given IBD levels (0.01, 0.05, 0.1, 0.15, 0.20, 0.25, 0.5, 0.75 and 0.99) in the Bhutanese national (n=12) and non-national (n=15) populations. At the data-generating r (IBD) of 0.5, the RMSE is approximately two-fold higher for the microsatellites than the microhaplotypes in both populations. Panel b) presents IBD-based cluster plots for each marker panel at minimum 3% relatedness using the Bhutanese sample set (12 national and 15 non-national cases). Greater distinction of non-national from national cases is observed with the microhaplotypes than the microsatellites.

## Discussion

Our study presents novel genetic data that serve as a foundation for the molecular surveillance of *P. vivax* in South Asia. Our high-resolution genetic data were derived from parasite isolates collected across multiple countries in the region, enabling a detailed investigation of *P. vivax* cross-border transmission and highlighting the risks for malaria resurgence in pre-elimination countries. The study also demonstrates the power of microhaplotype genotyping to distinguish local from cross-border importations.

As demonstrated in other studies, genetic diversity indices can be a useful tool in gauging the intensity of *P. vivax* transmission [28-31]. Our genetic data align with epidemiological data on annual parasite incidence (API), suggesting low transmission in Bangladesh, Bhutan, and Nepal, characterised by the absence of polyclonal infection and indicative of infrequent superinfection. The absence of polyclonal infections is not a limitation of the marker diversity, as this was high in all populations assessed. Conversely, there were high levels of polyclonal infection in Afghanistan and Pakistan, where API was high.

While there was a clear trend between within-host diversity and API, notable differences in population-level genetic relatedness were observed between countries. Samples from Bangladesh exhibited high relatedness between infections, suggesting potential bottlenecking due to reduced transmission and inbreeding. In contrast, samples from Bhutan (even when restricted to isolates collected from nationals) and Nepal showed low relatedness between infections, which could reflect the modest sample sizes in these populations. Alternatively, the influx of imported cases may sustain local diversity in these populations. In Bhutan, the country-wide spatial and long temporal sampling catchment could contribute to the low relatedness between infections. The mountainous terrain in Bhutan may reduce human movement between different provinces, which could lead to a highly structured local parasite population. Our analyses revealed apparent relatedness among *P. vivax* populations within and across geographically proximate border regions. These patterns highlight that while local transmission persists within each country, cross-border movement of parasites also remains a contributor to ongoing transmission.

Previous genetic surveys in Nepal, Bhutan and Bangladesh have primarily employed panels of up to 10 microsatellite markers [7, 8, 32]. However, as our analysis in Bhutan demonstrated, small microsatellite panels have low-resolution information and are unable to measure IBD accurately, constraining distinction between local and imported cases in porous border regions. In contrast, the microhaplotype-based IBD analysis in Bhutan successfully distinguished locally acquired infections from those genetically linked to parasites from neighbouring countries. This highlights the utility of microhaplotype genotyping in improving the detection of cross-border importations.

In contrast to Bhutan, Nepal had limited evidence of within-country population structure or differentiation from highly endemic areas in Afghanistan, India and Pakistan. This may reflect high parasite diversity sustained by cross-border importations from neighbouring populations. Future work combining genetic with epidemiological data in model frameworks to reconstruct transmission chains, and conducting source–sink mapping, could provide even clearer insights into cross-border transmission dynamics in areas such as Nepal [33]. Future work should also use available high-resolution microhaplotype data from high-endemic countries to gain deeper insights into local heterogeneity and genetic diversity.

Targeted genotyping by sequencing is more affordable than whole-genome sequencing; however, practicalities such as economies of scale need to be considered in countries nearing elimination, where case numbers are low, but surveillance requirements remain high. The requirements for sequencing infrastructure, analytical capacity and turnaround times for decision-making need to be carefully considered. Portable technologies such as nanopore sequencing could improve surveillance by enabling faster, field-based genetic analysis, especially in remote areas. For surveillance purposes, the timing of feedback to national malaria programs should match the level of transmission and program needs. In Bhutan, for example, the malaria program recommends quarterly or half-yearly genomic reports to track trends, with additional analyses conducted when outbreaks or imported cases are suspected. Our study was limited by using retrospective sample collections and a relatively small number of samples from several countries. The years of sample collection and catchment areas also differed between studies. However, in Bhutan and Nepal, which had the smallest sample size, the main constraint was the local endemicity: although small, the samples represent a significant proportion of all national cases. Despite the limitations, the data provides important proof-of-concept and baseline data for future surveillance in the region.

## Conclusion

Our findings demonstrate that high-resolution *P. vivax* genotyping provides a feasible and cost-effective approach for tracking intervention efficacy and cross-border spread. Continued investment in genetic infrastructure, capacity building, and collaborative data sharing will be critical to fully utilise the potential of genomic epidemiology to inform timely public health responses that will accelerate malaria elimination in South Asia and beyond.

## Author contributions

S. Auburn and A. Rai were involved in the conception and design of the study. A. Rai and A. Rumaseb performed the lab experiments. E.S., A. Rai, H.T., and K.S.H performed data analysis. P.G., S.W., T.D., M.S.A., N.G., A.G.R., N.A., S. Adhikari, M.R.B., K.R.R., R.N., R.R., B.Q., S.Z., M.A.B., B.L., R.N.P., and K.T. provided critical clinical and metadata information and resources, and local epidemiological insight that were central to the study. A. Rai and S. Auburn wrote the original draft of the manuscript. All authors reviewed and contributed to the final manuscript.

## Acknowledgements

We thank the patients who contributed their samples to the study, and the health workers and field teams who assisted with the sample collections.

## Data availability

The genetic data generated in this study (Bangladesh, Bhutan and Nepal) is openly accessible in the European Nucleotide Archive (Project accession PRJEB105559; Supplementary Data 1). The comparator data generated in previous studies (from Afghanistan, Pakistan and India) is also available in the European Nucleotide Archive (Supplementary Data 1).

