## Supplementary Information for "Genetic evidence of cross-border *Plasmodium vivax* spread in a malaria pre-elimination region of South Asia"

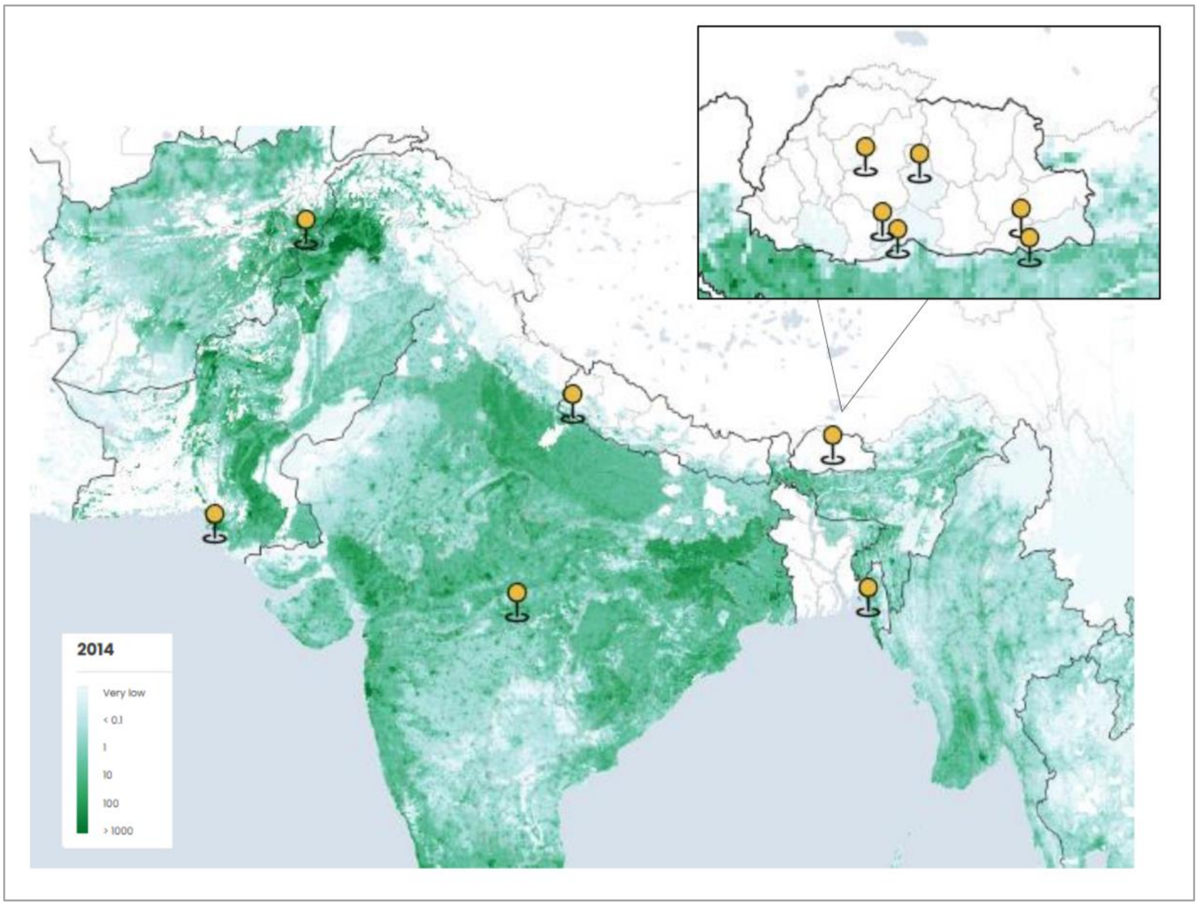


**Supplementary Figure 1. Geographic location of the study sites from which data were generated.** The large base map illustrates the countries where samples were derived from, including Nepal, Bhutan, Bangladesh, Afghanistan, Pakistan, and India. The smaller map illustrates the multiple divisions within Bhutan where samples were collected from. This map was generated using the Malaria Atlas Project tools and presents information on *P. vivax* clinical counts in 2014 (https://data.malariaatlas.org/maps).

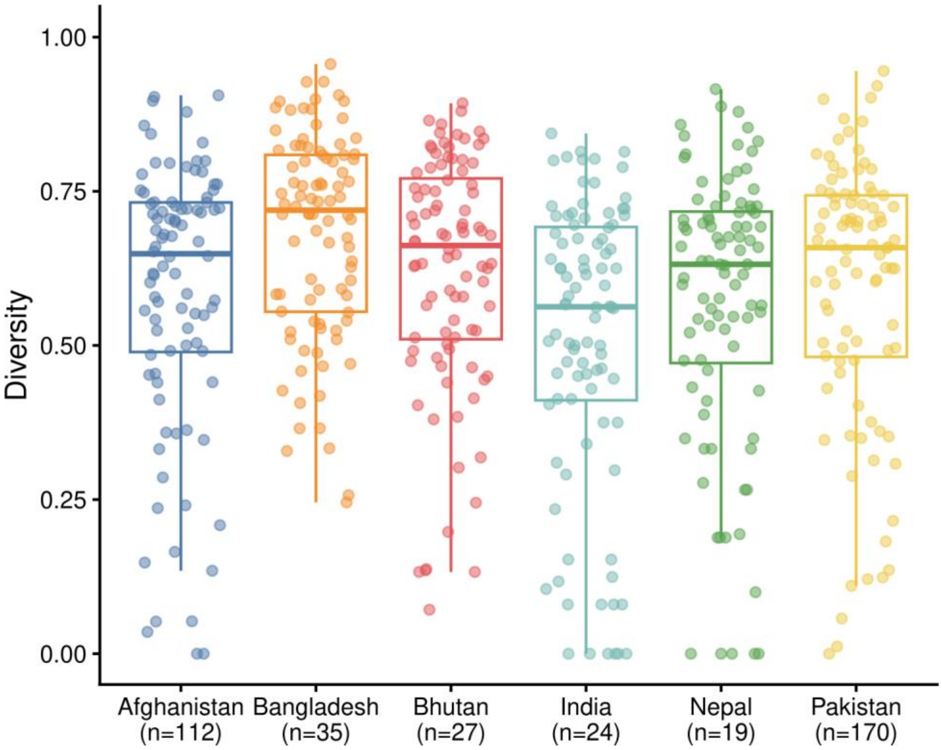


**Supplementary Figure 2. Marker diversity by country.** Microhaplotype marker diversity was analysed for samples from different countries using paneljudge [1]. Each box represents a country. Individual dots represent diversity estimates for each marker. Analysis was conducted on a total of 387 independent, monoclonal infections.

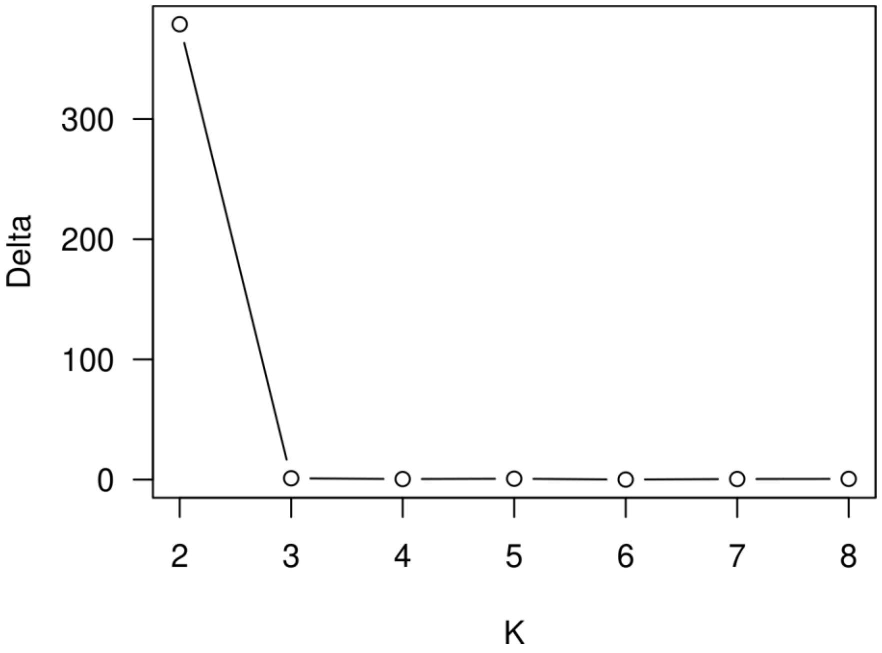


**Supplementary Figure 3. The optimal number of clusters (K).** The optimal number of clusters was determined to be K = 2, as calculated by CLUMPAK [2], as previously described [3].

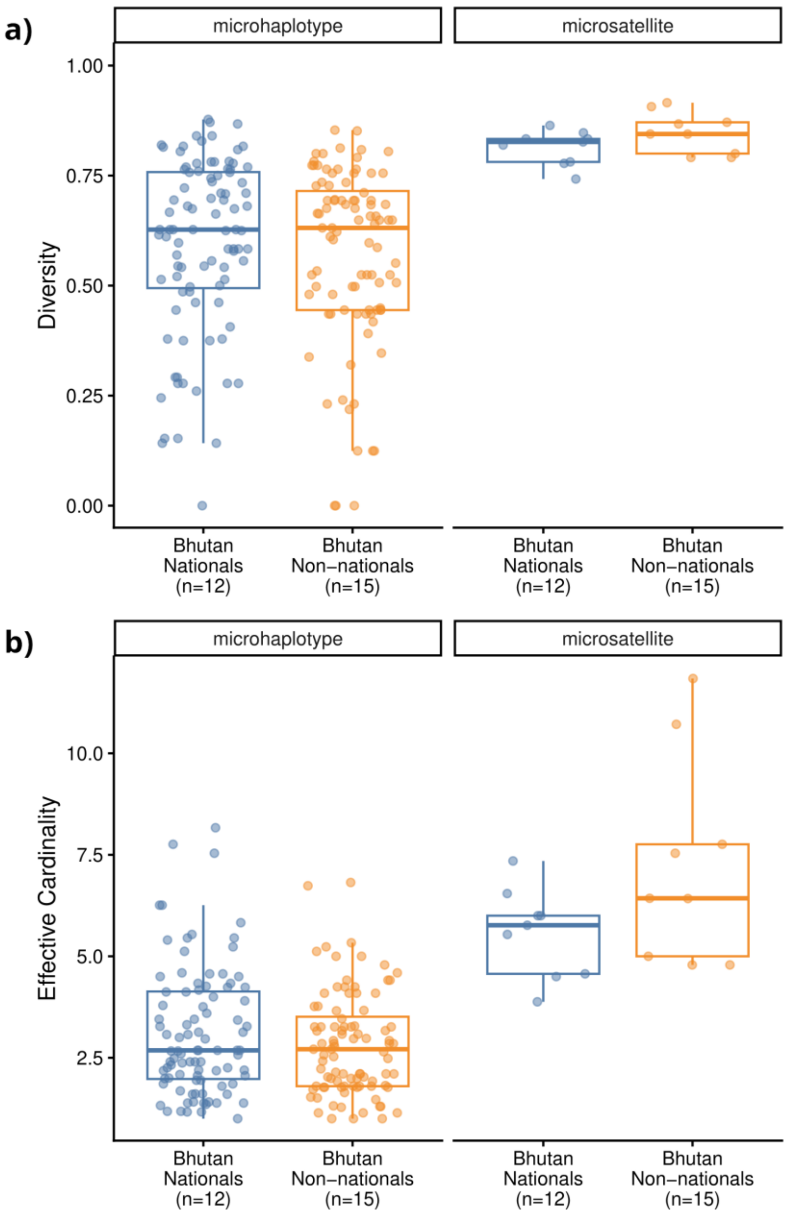


**Supplementary Figure 4. Marker diversity of the two marker panels.** Panel a) shows heterozygosity measures for Bhutan Nationals and Non-national cases, and panel b) shows effective cardinality scores in 12 independent Bhutan-National and 15 independent Bhutan Non-National samples. Each box plot shows the median, interquartile range, and minimum and maximum values for heterozygosity (panel a) and effective cardinality (panel b) calculated using paneljudge [1].

**Supplementary Table 1. Study site details**

| **Site name** | **Country** | **State/Division** | **Enrolment framework** | **Enrolment period** | **State/Province level *P. vivax* API 2014, 2024*** |
|---|---|---|---|---|---|
| Tikapur | Nepal | Sudurpaschim | Clinical trial | Aug 2021 - Aug 2023 | 0.68, 0.02 |
| Malakheti | Nepal | Sudurpaschim | Clinical trial | Aug 2021 - Aug 2023 | 0.68, 0.02 |
| Alikadam | Bangladesh | Chittagong Hill Tracts | Cross-sectional survey | Aug 2015 – Jan 2016 | 0.701.17 |
| Laghman | Afghanistan | Laghman | Clinical trial | July 2014 - Nov 2017 | 32.72, 32.06 |
| Jalalabad | Afghanistan | Nangarhar | Clinical trial | July 2014 - Nov 2017 | 48.03, 43.13 |
| Karachi | Pakistan | Sindh | Clinical trial | April 2023 – Sept 2024 | 9.83, 66.36 |
| Thatta | Pakistan | Sindh | Clinical trial | April 2023 – Sept 2024 | 9.83 66.36 |
| Sarpang tar/Rabdeyling /Jambeling/Ch ubarthang/Ch | Bhutan | Sarpang | Clinical efficacy | April 2013 – Oct 2015 | 0.54, 0 |

| | | | | | |
|---|---|---|---|---|---|
| okhorling/Umling | | | | | |
| Tsirang | Bhutan | Tsirang | Clinical efficacy | April 2013 – Oct 2015 | 0.1, 0 |
| Kamichu | Bhutan | Wangdiphodrang | Clinical efficacy | April 2013 – Oct 2015 | 0**, 0 |
| Sherubling | Bhutan | Trongsa | Clinical efficacy | April 2013 – Oct 2015 | 0.04, 0 |
| Mgalam | Bhutan | Pemagatshel | Clinical efficacy | April 2013 – Oct 2015 | 0**, 0 |
| Sethi | Bhutan | Samdrup Jongkhar | Clinical efficacy | April 2013 – Oct 2015 | 0**, 0 |

*API, Annual Parasite Incidence (cases/1000) for 2014 and 2024, derived from Malaria Atlas Project. **<0.001

**Supplementary Table 2. Demographic details of the patient samples that were successfully genotyped**

| Site | Country | Collection period | Age category (years) | | | % Male patients |
|---|---|---|---|---|---|---|
| | | | <5 | 5-15 | >15 | |
| Tikapur | Nepal | Oct 2021 - Aug 2023 | 0/14 (0%) | 0/14 (0%) | 14/14 (100%) | 85.7% (12/14) |
| Malakheti | Nepal | Oct 2021 - Aug 2023 | 0/5 (0%) | 1/5 (20%) | 4/5 (80%) | 80.0% (4/5) |
| Bhutan | Bhutan | May 2013 - Apr 2015 | 0/27 (0%) | 3/27 (11.1%) | 24/27 (88.9%) | 88.9% (24/27) |
| Chittagong Hill Tracts | Bangladesh | Aug 2014 - Jan 2015 | 2/35 (5.7%) | 12/35 (34.3%) | 21/35 (60%) | 60.0 % (21/35) |
| Laghman | Afghanistan | Sep 2014 - Apr 2016 | 3/30 (10%) | 22/30 (73.3%) | 5/30 (16.7%) | 66.7% (20/30) |
| Jalalabad | Afghanistan | Apr 2015 - Apr 2016 | 13/129 (10.1%) | 53/129 (41.1%) | 63/129 (48.8%) | 75.2 % (97/129) |
| Karachi | Pakistan | May 2023 - Mar 2024 | 0/22 (0%) | 0/22 (0%) | 22/22 (100%) | 72.7% (16/22) |
| Thatta | Pakistan | Apr 2023 - Mar 2024 | 0/191 (0%) | 0/191 (0%) | 191/191 (100%) | 73.3% (140/191) |

**Supplementary Table 3. List of Ethics Approvals**

| Country | Ethics review board/regulatory authorities | Reference number |
|---|---|---|
| Australia/Nepal | Human Research Ethics Committee of the Northern Territory Department of Health and Menzies School of Health Research | HREC-2019-3467 |
| | Nepal Health Research Council | 857/2019 P |
| Australia/Bhutan | Human Research Ethics Committee of the Northern Territory Department of Health and Menzies School of Health Research | HREC-2012-1871 |
| | Research Ethics Board of Health, at the Ministry of Health in Bhutan | REBH 2012/031 |
| Australia/Bangladesh | Human Research Ethics Committee of the Northern Territory Department of Health and Menzies School of Health Research | HREC-2014-2228 |
| | Ethics Review Committee of the icddr,b | PR-14053 |

| | | |
|---|---|---|
| Australia/Afghanistan | Human Research Ethics Committee of the Northern Territory Department of Health and Menzies School of Health Research | 13–1991 |
| | Oxford Tropical Research Ethics Committee OxTREC | 1014–13 |
| | Ministry of Public Health, Afghanistan | |
| Australia/Pakistan | Human Research Ethics Committee of the Northern Territory Department of Health and Menzies School of Health Research | HREC-20-3694 |
| | Ethics Review Committee of the Aga Khan University | 2022-7042-20769 |
| | National Institutes of Health, Health Research Institute, National Bioethics Committee (NBC) | 4-87/NBC-772/22/1833 |
| | Drug Regulatory Authority of Pakistan | 16-37/2022 DD (PS) |
